# Identification of candidate genes for developmental colour agnosia in a single unique family

**DOI:** 10.1101/2021.10.01.21263387

**Authors:** Tanja C. Nijboer, Ellen V.S. Hessel, Gijs W. van Haaften, Martine J. van Zandvoort, Peter J. van der Spek, Christine Troelstra, Carolien de Kovel, Bobby P.C. Koeleman, Bert van der Zwaag, Eva H. Brilstra, J. Peter H. Burbach

**Affiliations:** UMCU Brain Center and Center of Excellence for Rehabilitation Medicine, University Medical Center Utrecht and De Hoogstraat Rehabilitation, 3584 CX Utrecht, The Netherlands; Department of Experimental Psychology and Helmholtz Institute, Utrecht University, 3584 CS Utrecht, The Netherlands; Department of Biomedical Genetics, University Medical Center Utrecht, 3584 CG Utrecht, The Netherlands; UMCU Brain Center, Department of Translational Neuroscience, University Medical Center Utrecht, 3584 CG Utrecht, the Netherlands; Department of Pathology, Erasmus Medical Center Rotterdam, 3015 GD Rotterdam, the Netherlands

**Author notes:** These authors contributed equally (Tanja C. Nijboer, Ellen V.S. Hessel, Gijs W. van Haaften).

## Abstract

Colour agnosia is a disorder that impairs colour knowledge (naming, recognition) despite intact colour perception. Previously, we have identified the first and only-known family with hereditary developmental colour agnosia. The aim of the current study was to explore genomic regions and candidate genes that potentially cause this trait in this family. For three family members with developmental colour agnosia and three unaffected family members CGH-array analysis and exome sequencing was performed, and linkage analysis was carried out using DominantMapper, resulting in the identification of 19 cosegregating chromosomal regions. Whole exome sequencing resulted in 11 rare coding variants present in all affected family members with developmental colour agnosia and absent in unaffected members. These variants affected genes that have been implicated in neural processes and functions (*CACNA2D4, DDX25, GRINA, MYO15A*), that have a indirect link to brain function or development (*MAML2, STAU1, TMED3*), and a remaining group lacking brain expression or involved in non-neural traits (*DEPDC7, OR1J1, OR8D4, RABEPK*). Although this is an explorative study, the small set of candidate genes that could serve as a starting point for unravelling mechanisms of higher level cognitive functions and cortical specialization, and disorders therein such as developmental colour agnosia.

## Introduction

Genetics of developmental disorders of the human brain has been a main strategy to uncover biological mechanisms of brain development and functioning. In particular, understanding of the development of the cerebral cortex has benefited from the recognition of specific cortical abnormalities and the elucidation of causative gene defects (Ayala, Shu, & Tsai, 2007; Pilz et al., 1998; Sossey-Alaoui et al., 1998). Genetic aberrations in cortical specializations may be a treasure trove to find genetic dominators and mechanisms of cognitive functions (Hu, Chahrour, & Walsh, 2014; Mitchell, 2011).

Different disorders of cortical specialization have been described (reviewed by(Mitchell, 2011): prosopagnosia (i.e. inability to recognise faces/people despite normal intelligence), synesthesia (i.e. stimulation of one sensory or cognitive pathway leads to automatic, involuntary experiences in a second sensory or cognitive pathway, for example, letters or numbers are perceived as inherently coloured), dyslexia (i.e. disability in reading despite normal intelligence), and congenital amusia (i.e. defect in processing pitch, but may also include musical memory and recognition).

In this study we focussed on a very rare disorder of cortical specialization, namely developmental colour agnosia. Disorders of colour processing have been reported at different levels, ranging from wavelength processing deficits up to very specific impairments in object-colour associations (De Vreese, 1991). One of the most intriguing disorders may be colour agnosia. In colour agnosia, people have intact colour perception, yet have severe difficulties in naming, categorising, and recognising colours and form adequate object-colour associations (Nijboer, van Zandvoort, & de Haan, 2006, 2007a, 2007b; van Zandvoort, Nijboer, & de Haan, 2007). In most cases, colour agnosia is acquired after brain damage, either bilateral or left hemisphere lesions (De Renzi, 2000), mostly in the occipitotemporal lobe (De Vreese, 1991).

Over a decade ago we reported the first and only case of developmental colour agnosia (van Zandvoort et al., 2007). Additionally, a familial factor in developmental colour agnosia was described when family members in three generations also showed difficulties in colour naming and recognition (Nijboer, van Zandvoort, et al., 2007a), indicating a genetic origin of the trait. The affected individuals of this family all display normal cognition and intact colour perception at a visuo-sensory level, e.g. matching of equiluminant colours. However, they could not name colours, match colour names to colours, categorise hues into general clusters of colour or point a colour that is mentioned by the examiner (Nijboer, van Der Smagt, van Zandvoort, & De Haan, 2007). This trait likely is a disorder in cortical specialization (Mitchell, 2011). Such disorders may have a genetic origin resulting in abnormal cortical development.

In order to identify genetic origins and to gain more insight into the developmental mechanisms related to these disorders, we analyzed all participating members of this family by CGH-arrays and whole-exome sequencing. The results reveal a small set of extremely rare variants that are candidate to cause developmental colour agnosia within the limits of this study.

## Subjects and Methods

### Subjects and DNA isolation

Informed and written consent were obtained from the family members to perform DNA analysis and sequencing, and to publish results in a scientific paper as approved by the Medical Ethical Committee of the University Medical Center Utrecht. Four members were phenotyped earlier (Nijboer, van Zandvoort, et al., 2007a). For this study, two additional family members were tested and diagnosed with or without developmental colour agnosia. Phenotyping has been described by (Nijboer, van Der Smagt, et al., 2007; Nijboer, van Zandvoort, et al., 2007a). In total three affected and three unaffected family members were available for this study (Fig. 1). Genomic DNA of the six family members was isolated from saliva according to standard procedures. The DNA concentration was determined by using Qubit Quant-IT (Invitrogen).

**Fig. 1.**
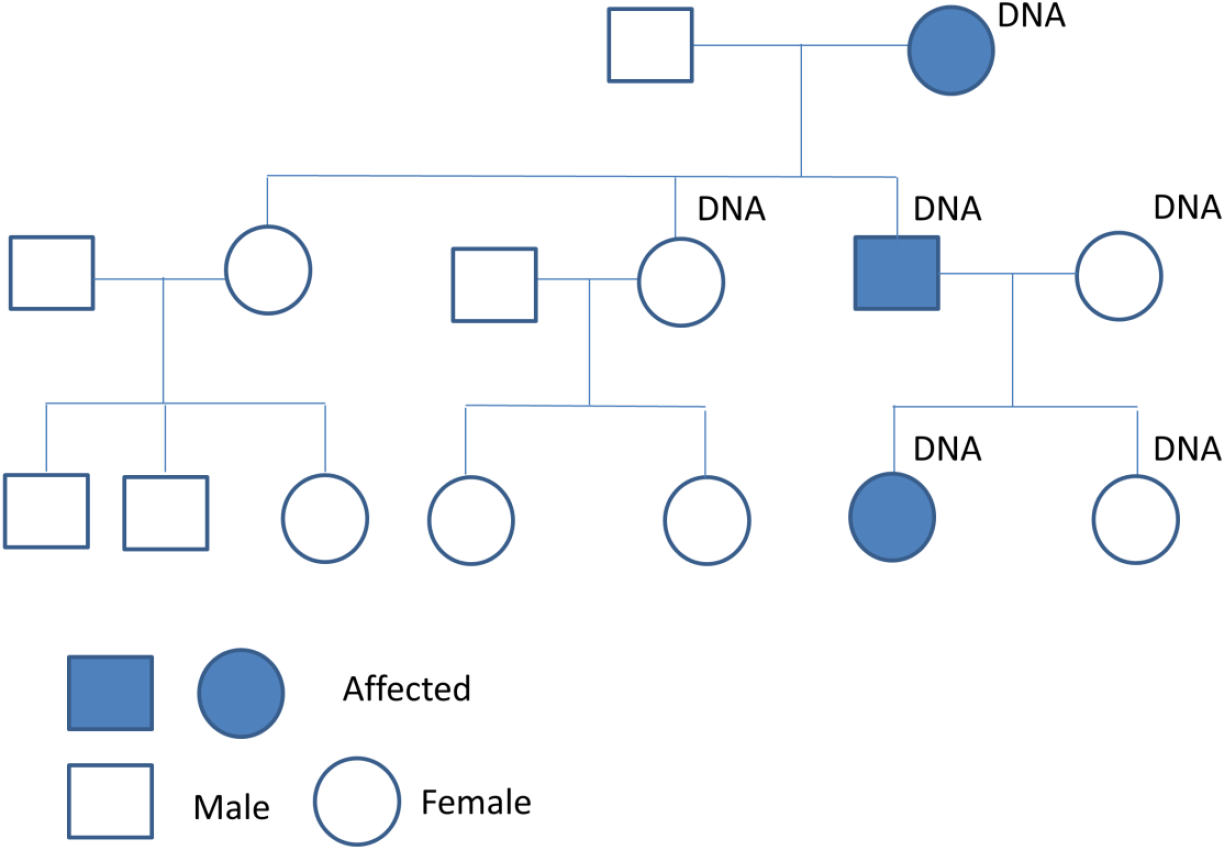
Family with Developmental Colour Agnosia. DNA means that these family members underwent genetic analysis. Three affected persons are indicated (filled dark blue).

### Array-based comparative genomic hybridization and Linkage Analysis

Array-CGH microarray analysis was performed using 105K (Agilent amadid 019015, hg18) microarray slides from Agilent Technologies (Santa Clara, CA) following manufacturer’s protocols and a mixed pool of either 50 healthy males or females was used as a reference. Scanned images were analyzed by Feature Extraction software (Agilent Technologies). Data analysis was performed using DNA analytics 4.76 Software from Agilent Technologies using the ADM-2 algorithm. Interpretation of CNV-data was performed as described in the guidelines of (Vermeesch et al., 2007). Copynumber variations and sequence homologies flanking the breakpoints were confirmed using the Database of Genomic Variation (http://www.projects.tcag.ca/variation/) and the UCSC Genome Browser (http://www.genome.ucsc.edu/index) (Van Binsbergen, Hochstenbach, Giltay, & Swinkels, 2012).

Linkage analysis was performed by using the array CGH chromosome profiles and mapping of the disease loci was calculated with DominantMapper (Carr et al., 2011). This program calculated which chromosomal regions are present in all affected family members and not present in all non-affected family members based on SNP genotyping data.

### Library preparation and whole exome Sequencing

DNA (2ug) was used for the preparation of a barcoded fragment library (Harakalova et al., 2012). This was followed by multiplexed enrichment and sequencing on the SOliD Next-generation sequencing platform containing the exonic sequences of ∼18.000 genes and covering a total of ∼37Mb of genomic sequence (Harakalova et al., 2011, 2012). In brief, the DNA was fragmented using the Covaris S2 System (Applied Biosystems), after fragmentation, the DNA was end repaired and phosphorylated at the 5’s end using the End-It DNA End-Repair Kit (EpiCenter) and purified with the Agencourt AMPure XP system (Beckman Coulter Genomics). DNA was then ligated to double-stranded truncated adaptors compatible with the SOLiD next-generation sequencing platform using the Quick Ligation Kit (NEB). After purification, each sequencing library was nick translated, barcoded and amplified in a single PCR assay. The intensity of library bands was examined on a 2% agarose gel (Lonza FlashGel System). Amplified library fragments in the range of 175–225 bp in size were selected on a 4% agarose gel and purified using a QIAquick Gel Extraction Kit (Qiagen). Libraries from the family members were pooled in equimolar concentrations and were enriched using the Agilent SureSelect Human All Exon 50Mb Kit (Agilent Technologies). Enriched library pool fragments were amplified using 12 PCR cycles and elongated to a full-length adaptor sequence required for SOLiD sequencing. SOLiD sequencing was performed according to the instructions in the SOLiD 4 manual to produce enough 50-bp reads to obtain sufficient coverage for a single allele in each library.

### V***ariant detection and exome sequencing analysis***

Raw sequencing reads were mapped against the human reference genome GRCh37/hg19 using our custom pipeline based on the Burrows-Wheeler Alignment (BWA) algorithm26. Sequence data were submitted to the EMBL-EBI Sequence Read Archive (see URLs). Single-nucleotide variants (SNVs) and small indels (≤7 nt) were called by our custom analysis pipeline as described (Nijman et al., 2010) (all scripts are available upon request). The criteria for variant detection were set to enable discovery of heterozygous variants, since we assume dominant inheritance. We required that variants should minimally be supported by two read seeds (first 25 bp, the higher quality portion of a read), and we set the cut-off for coverage to a minimum of ten reads and the cutoff for non-reference allele percentage to 15%. Since it is probably a dominant inherited phenotype, we set the cut-off for non-reference allele percentage to a maximum of 80%. A maximum of five clonal reads (defined as reads with an identical start site) were included in the analysis. Only truncating variants including nonsense frameshifting insertions or deletions, or missense and splice site alterations were selected. All polymorphisms with an allele frequency < 0.002 in the different databases in EVS database (http://evs.gs.washington.edu/EVS/), Exac (http://exac.broadinstitute.org/) or GoNL (http://www.nlgenome.nl/) were excluded, and variants present in our in-house database > 0,1,2 x present were excluded from analysis; For each variant, the genomic location, amino-acid change, effect on protein function, conservation score and output from prediction programs (Polyphen, Polyphen-2, SIFT and Condel) were compiled (Harakalova et al., 2012). Alamut was used to study the variants in more detail. Prioritization and variant selection is described in Fig. 2.. Identified variants were confirmed with Sanger sequencing.

**Fig. 2.**
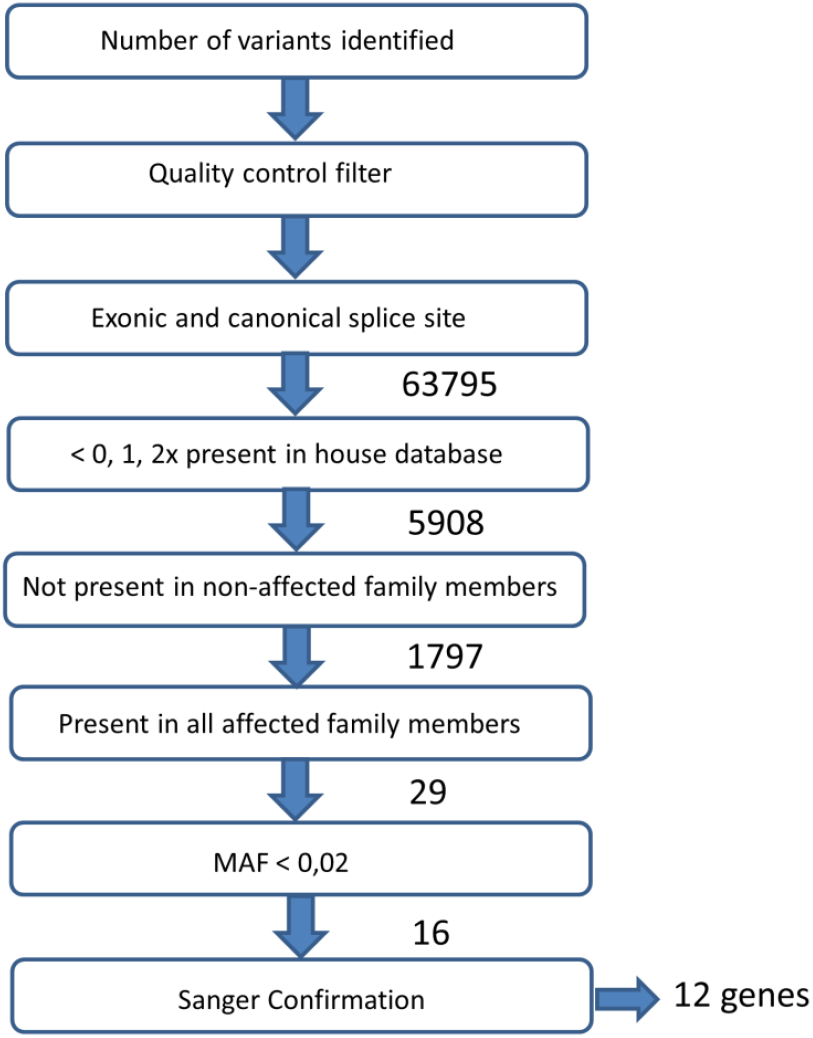
Selection of variants cosegregating with developmental colour agnosia based on whole exome sequencing. Selection is based on the presence of the mutation in the affected family members and the absence in the unaffected family members.

### Gene expression analyses

Comprehensive human brain transcriptome data sets were used to determine the developmental expression profiles of all candidate genes (Visser et al., 2010). The set contains the age-related expression in 39 samples of the dorsolateral prefrontal cortex from individuals ranging from 0.1 to 83 years of age (Gene Logic Inc, Gaithersburg, MD, USA ; (Stubbs et al., 2012; Visser et al., 2010). Raw data of these studies were available online as GEO entry (GSE11882, Figure 3).

**Fig. 3.**
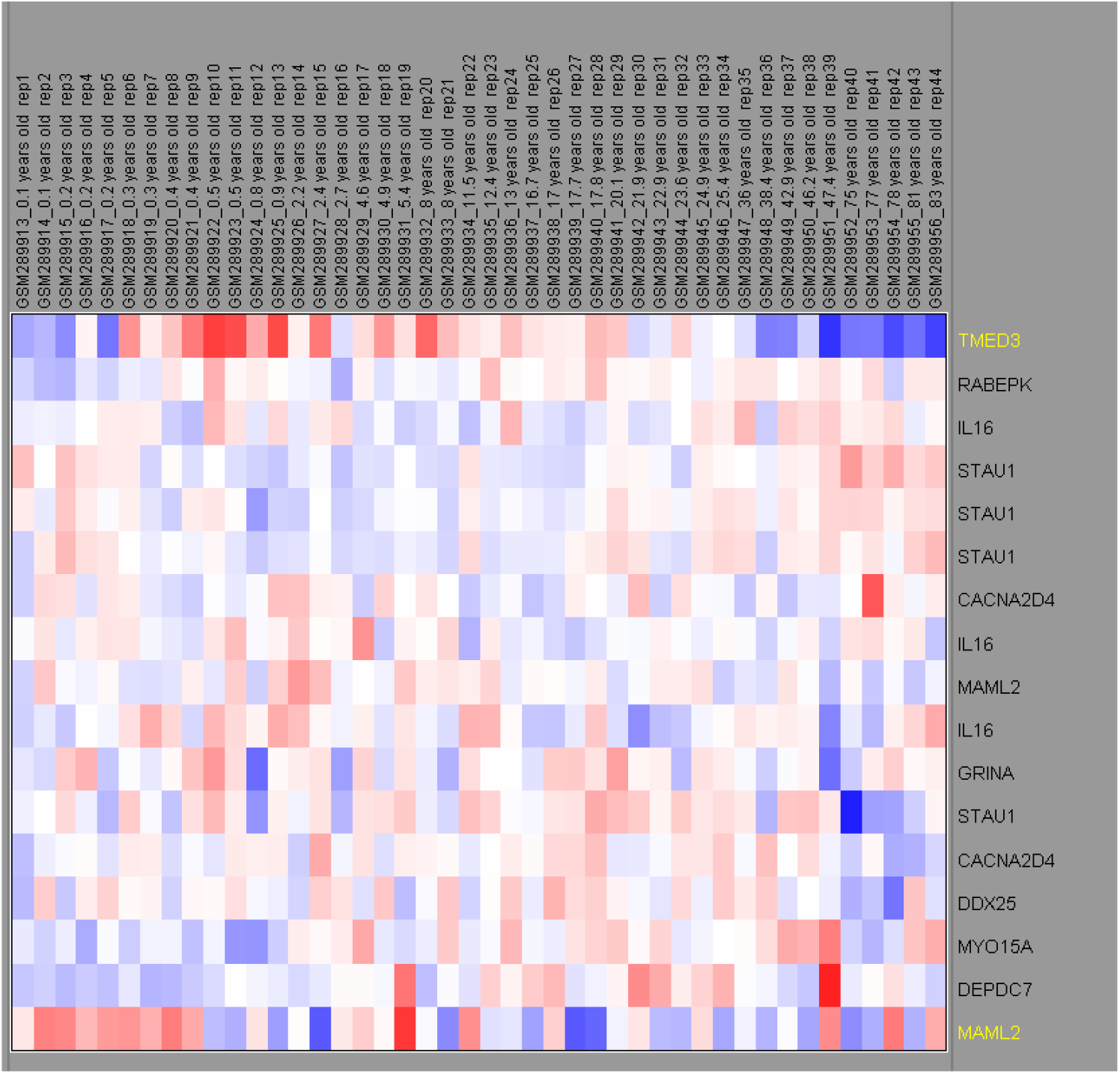
Expression of the candidate genes in the human brain (prefrontal cortex) during development. The upper lane describes the age of the brain samples and sample ID. The gene names are mentioned at the right. Red box means an up regulation, blue box is a down regulation and white is no change in expression during development.

For analysis of gene expression single-cell RNAseq datasets provided by the Allen Brain Institute were used in the Allen Brain Map Transcriptomics Explorer (https://portal.brain-map.org). For the human brain on datasets Human-M1-10X and Human-Multiple cortical areas-SMART-seq were used (Hodge et al., 2019). For the mouse brain the data set Mouse-Whole mouse brain & hippocampus-10X was used. In addition, data of the mouse cortex (Tasic, Menon, Nguyen, Kim, Jarsky, Yao, Levi, Gray, Sorensen, Dolbeare, Bertagnolli, Goldy, Shapovalova, Parry, Lee, Smith, Bernard, Madisen, Sunkin, Hawrylycz, Koch, Zeng, et al., 2016; Tasic et al., 2018) were mined through the Single Cell Portal (https://singlecell.broadinstitute.org/single_cell) and the Allen Brain Atlas data portal (http://casestudies.brain-map.org/celltax#section_explorea). The Mouse Brain Atlas and Developing Mouse Brain Atlas were used for anatomical analysis of gene expression (https://portal.brain-map.org).

## Results and Discussion

### CGH-array and Linkage Analysis

Segregration patterns of developmental colour agnosia in the family are most consistent with Mendelian dominant inheritance (Fig 1). Array CGH performed on the six family members showed that there are no large chromosomal rearrangements co-segregating with the phenotype of developmental colour agnosia. Next, we performed linkage analysis on SNP data to identify loci cosegregating with developmental colour agnosia. All family members were included in the linkage analysis assuming an autosomal dominant model of inheritance. The genome wide linkage resulted in the identification of multiple linked chromosomal regions (Table 2). These linked regions were used for the identification of the candidate genes assuming that the causal gene will be present in a linked region.

**Table 1:** Linked chromosomal regions to colour agnosia.

| Chr. | Locus | Start | End | Length |
| --- | --- | --- | --- | --- |
| 2 | 2q22.3-35 | 146360626 | 221800201 | 75439575 |
| 3 | 3p11.2-q11.2 | 87373894 | 94512868 | 7138974 |
| 3 | 3q13.3-q25.31 | 119442572 | 159239866 | 39797294 |
| 4 | 4q13.1-4q22.3 | 60013919 | 96704454 | 36690535 |
| 5 | 5p15.33-5p15.2 | 2346621 | 10752315 | 8405694 |
| 6 | 6q26-27 | 162736476 | 168554222 | 5817746 |
| 8 | 8q24.23-24.3 | 138970102 | 143270729 | 4300627 |
| 8 | 8p22-q13.1 | 14253354 | 68723609 | 54470255 |
| 9 | 9q21.32-q34.11 | 83858558 | 138149166 | 54290608 |
| 10 | 10p15.3-p14 | 135708 | 8638235 | 8502527 |
| 11 | 11p14.3-q24 | 21234023 | 128206410 | 106972387 |
| 12 | 12p13-q23.3 | 661031 | 67262352 | 66601321 |
| 14 | 14q12-q31.2 | 22645683 | 88752821 | 66107138 |
| 15 | 15q15.1-q26.1 | 40509600 | 94077313 | 53567713 |
| 17 | 17p12-q25.3 | 9078327 | 77698582 | 68620255 |
| 19 | 19p12-q21 | 23446279 | 28836326 | 5390047 |
| 20 | 20q13.13-q13.33 | 47050483 | 60111743 | 13061260 |
| 21 | 21q11.2-q22.12 | 14687571 | 38082812 | 23395241 |
| 22 | 22q11.2-q13.2 | 16079545 | 44595591 | 28516046 |

Chromosomal regions linked to developmental colour agnosia (hg19). These haplotypes of these regions are present in all affected individuals and absent in the non-affected individuals.

**Table 2:** Properties of selected candidate genes.

| Genome position<br>(hg19) | Nucleotide change | Gene symbol | Ensemble transcript | Consequence | Amino acid change | SNP id | Protein function |
| --- | --- | --- | --- | --- | --- | --- | --- |
| 8_145066215 | G/A | GRINA | ENST00000313269 | NON_SYNONYMOUS | R221Q | novel | Glutamate Receptor |
| 9_125239752 | C/T | OR1J1 | ENST00000259357 | NON_SYNONYMOUS | A152T | rs143862742 | Olfactory receptor |
| 9_127990266 | G/A | RABEPK | ENST00000373538 | NON_SYNONYMOUS | G202R | rs142347258 | RAB9 effector protein |
| 11_33052944 | C/T | DEPDC7 | ENST00000241051 | NON_SYNONYMOUS | A268V | novel | G protein signalling DEP domain |
| 11_95712527 | T/C | MAML2 | ENST00000524717 | NON_SYNONYMOUS | Q1019R | rs201299551 | Transcriptional Coactivator Notch |
| 11_123777296 | G/A | OR8D4 | ENST00000321355 | NON_SYNONYMOUS | R53H | rs201007236 | Olfactory receptor |
| 11_125786960 | C/G | DDX25 | ENST00000263576 | NON_SYNONYMOUS | D284E | rs370049057 | DEAD box RNA helicase |
| 12_1949897 | G/A | CACNA2D4 | ENST00000382722 | SPLICE_SITE | NA | rs376597005 | Calcium Channel |
| 15_79614380 | C/T | TMED3 | ENST00000299705 | NON_SYNONYMOUS | H160Y | novel | Transmembrane protein |
| 17_18043906 | C/T | MYO15A | ENST00000205890 | NON_SYNONYMOUS | R1763W | rs200146361 | Actin based motor molecule |
| 20_47740944 | T/C | STAU1 | ENST00000371856 | NON_SYNONYMOUS | R264G | rs201180807 | RNA binding protein |

### Exome sequencing and candidate gene selection

We hypothesized that developmental colour agnosia is caused by a single heterozygous rare non-synonymous variant present in all affected family members and absent in unaffected family members. Candidate gene selection was performed by filtering as described in Fig 2. Exome sequencing followed by quality control identified in total in all family members 63795 variants present in exons and canonical splice sites. Of these, 5908 variants were protein truncating with putative loss of function alteration, including nonsense frameshift (indels) or splice site variants, and were *de novo* or rare in the population (0, 1, 2 times present in the in-house database). Of these 5908 variants, 1797 were not present in non-affected family members and 29 were present in all three affected family members. Of these 29 variants we further studied the frequency of these variants, to assume that the variants are rare or *de novo* in a broader population. Based on frequency of presence in the Exome Variant Server (EVS) database (frequency 0:13000-25:13000 (frequency < 0.002)) (n=17) and Variance GoNL database (n=750; Francioli et al., 2014) (n=16), and confirmation of the variant in the linked region by Sanger sequencing (n=11), 11 candidate variants in 11 genes remained (table 2): *GRINA* (glutamate receptor, ionotropic, N-methyl D-aspartate-associated protein 1), *OR1J1* (olfactory receptor, family 1, subfamily J, member 1), *RABEPK* (Rab9 effector protein with Kelch motifs), *DEPDC7* (DEP domain containing 7), *MAML2* (mastermind-like 2 (Drosophila)), *OR8D4* (olfactory receptor, family 8, subfamily D, member 4), *DDX25* (DEAD (Asp-Glu-Ala-Asp) box polypeptide 25), *CACNA2D4* (calcium channel, voltage-dependent, alpha 2/delta subunit 4), *TMED3* (transmembrane emp24 protein transport domain containing 3), *IL16* (interleukin 16), *MYO15A* (myosin XVA), *STAU1* (staufen double-stranded RNA binding protein 1). Additional information including allele frequency in the 1000 Genome Project (Altshuler et al., 2010)(The 1000 Genomes Project Consortium), Variance GoNL (Boomsma et al., 2014; Francioli et al., 2014) and Wellderly cohort (Erikson, Deshpande, Kesavan, & Torkamani, 2015) is presented in Table 3.

**Table 3:** Variant frequencies and mutation prediction.

| Gene_name | Frequencies |  |  | Effect of mutation |  |  |  |  | Conservation (Condel) |  |
| --- | --- | --- | --- | --- | --- | --- | --- | --- | --- | --- |
|  | EVS | GoNL | Exac | Effect | Polyphen_s | Polyphen_p | SIFT_s | SIFT_p | Conserved Nucl. | Conserved AA |
| *GRINA* | 1/13005 | 0 | 0/121000 | NS | 1 | probably damaging | 0 | deleterious | highly | highly |
| *OR1J1* | 0 | 0 | 1/120930 | NS | 0,001 | benign | 0,56 | tolerated | weakly | highly |
| *RABEPK* | 11/12995 | 2/996 | 36/120704 | NS | 1 | probably damaging | 0 | deleterious | highly | highly |
| *DEPDC7* | 0 | 0 | 0/120000 | NS | 0,896 | possibly damaging | 0,07 | tolerated | moderately | moderately |
| *MAML2* | 1/12217 | 0 | 50/119130 | NS | 0,998 | probably damaging | 0,06 | tolerated | moderately | highly |
| *OR8D4* | 6/12996 | 1/996 | 102/121352 | NS | 0 | benign | 0 | deleterious | weakly | weakly |
| *DDX25* | 1/12537 | 1/996 | 3/76840 | NS | 0 | benign | 0,49 | tolerated | weakly | highly |
| *CACNA2D4* | 0 | 0 | 3/120548 | SS | undef | undef | undef | undef | na (splicesite) | na (splicesite) |
| *TMED3* | 0 | 0 | 0/121000 | NS | 0,773 | possibly damaging | 0,06 | tolerated | moderately | highly |
| *MYO15A* | 17/12365 | 4/996 | 199/119810 | NS | 0,999 | probably damaging | undef | undef | weakly | highly |
| *STAU1* | 3/13003 | 0 | 16/121402 | NS | 0,911 | possibly damaging | 0,33 | tolerated | weakly | highly |

Candidate variants and their frequency in different databases. Polyphen and SIFT predict the effect of the mutation. Effect NS is non-synonymous, SS splice site. Conserved Nucl. shows the conservation of the nucleotides and conserved AA determined the conservation of the AA.

### Effect of the mutation

Prediction about the effect of the mutations were made by three software packages: Polyphen, Polyphen-2, SIFT and Condel (Table 2). Two variants (*GRINA* and *RABEPK*) were predicted to be probably damaging (Polyphen) and deletious (SIFT). Six variants (*OR1J1, DEPDC7, MAML2, DDX25, TMED3, STAU1*) were predicted as tolerated by SIFT; some of these were benign or possibly damaging by Polyphen (*OR1J1, DDX25, DEPDC7, TMED3, STAU1*). *MAML2* was predicted as probably damaging by Polyphen. *OR8D4* was deleterious on SIFT and benign on Polyphen. MYO15A was probably damaging by Polyphen and undefined by SIFT. The effects of two variants (*IL16* and *CACNA2D4)* could not be predicted, since these are splice site mutations (Table 3).

### Gene Expression during human brain development

We have attempted to prioritize the 11 candidate genes for a possible role in developmental colour agnosia. For all these genes, we have analyzed (developmental) brain expression (Fig 3). We hypothesised cortical expression during development for a gene linked to developmental colour agnosia and possible increased expression during postnatal brain maturation. For that reason we first studied expression of the 11 candidate genes in the human brain of subjects free of neurological disorders, in the age range of 0.1 to 83 years, represented by the Ingenuity data set. All genes seem to be expressed in the brain, while two genes, *TMED3* and *MAML2*, displayed an increased expression during development (*TMED3*, 0.4-2.4 years old and *MAML2*, 0.1-2.4; Fig 3). The other nine genes do not show a clear expression profile during a specific developmental age window.

Mining of single-cell transcriptomic data of the adult human cortex and mouse brain showed that MAML2 was the only gene that maintained expression at high levels, albeit restricted to only a limited number of cell types (Fig 4). The other genes displayed a low to very low level of expression (*GRINA, RABEPK, DDX25, CACNA2D4, STA*U1) or no expression. In the human M1 motor cortex *MAML2* was expressed by three types of inhibitory interneurons, one type of excitatory neuron, and by no-neuronal cells: astrocytes, an olfactory progenitor neuron and microglia (Fig 4). In this respect it was de only gene that was expressed by non-neuronal cells. In the mouse cortex and hippocampus *Mam*l2 was also expressed in a very restricted set of neurons (dentate gyrus and CA3) and in non-neuronal cells. *Grina* and *Stau1* displayed wide, but low expression in inhibitory and excitatory neurons (Fig 4).

**Fig. 4.**
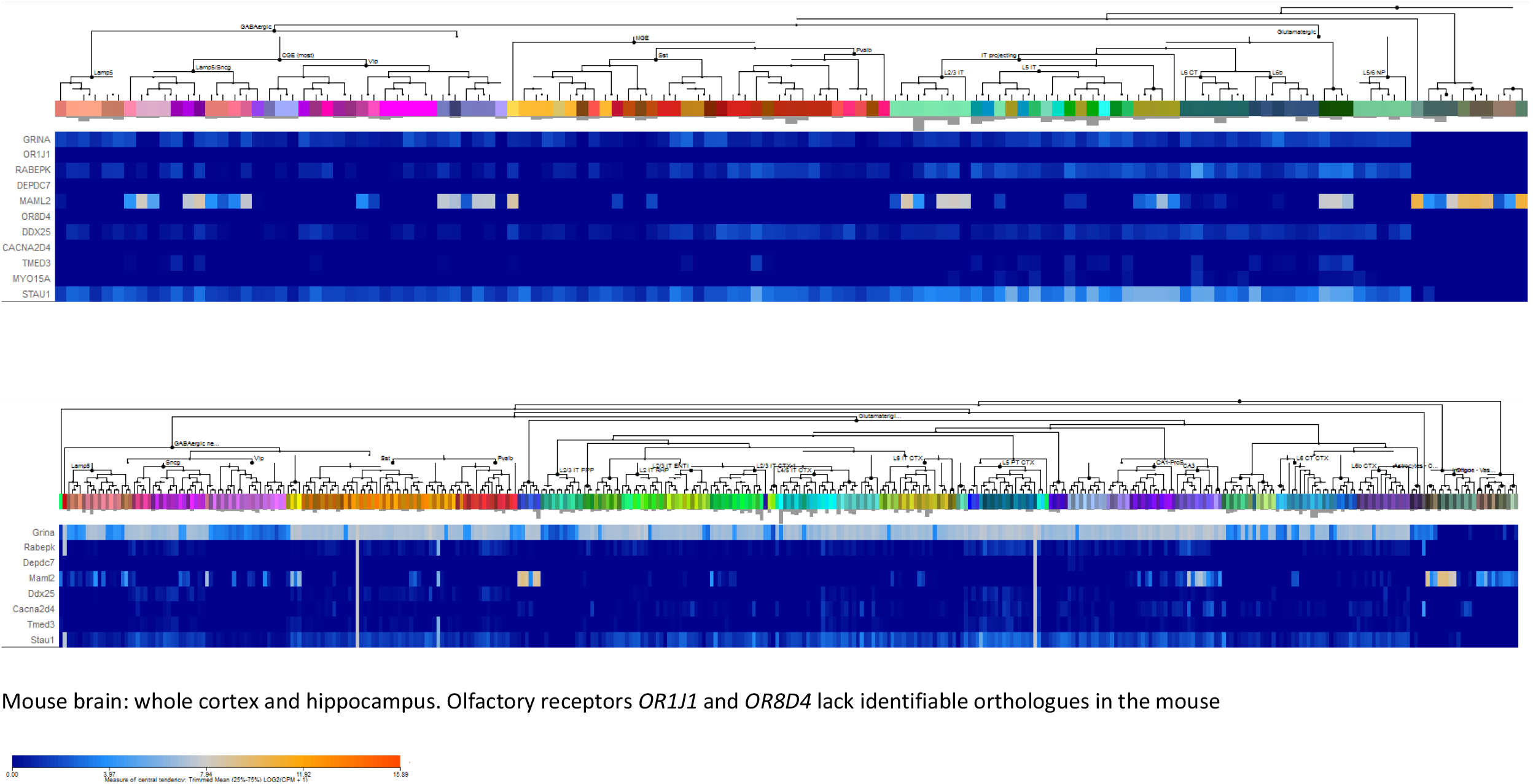
Expression of the candidate genes in different neuronal and non-neuronal cell types in the human brain (M1 motor cortex) and the mouse cortex and hippocampus. Data were obtained from the single-cell RNA-seq cell types databases of the Allen Brain Institute https://portal.brain-map.org/atlases-and-data/rnaseq). Expression levels are plotted under the neuronal and nonneuronal cell types identified in theses samples. Top: human, Bottom: mouse.

### Possible link of candidate genes to developmental colour agnosia

In an attempted to prioritize the 11 identified candidate genes for a possible role in colour agnosia, we have analysed gene function, link to visual perception, link to cortical specialization, conservation of the nucleotide or amino acid, frequency worldwide or in the Dutch population (Table 3) and its presence in previously linked regions for other cortical disorders for example prosopagnosia or dyslexia (Mitchell, 2011).

None of the identified genes or its variants was present amongst previously identified genes or loci related to dyslexia, dyscalculi or prosopagnosia. Linked regions were studied on OMIM (www.omim.org/). GO-analysis identified that the *calcium channel, voltage-dependent, alpha 2/delta subunit 4 (Cav1*.*4)* gene *CACNA2D4* and *MYO15A* were involved in visual perception, which makes *CACNA2D4 and MYO15A* candidate genes of interest.

*CACNA2D4* encodes the α2δ4 subunit of the photoreceptor voltage-gated calcium channels (Cav1.4), which modulate glutamate release from the photoreceptors to the bipolar cells. The CACNA2D4 mutation is a splice-site mutation and its role on the Cav1.4 protein function is unclear. Mutations in the human *CACNA2D4* gene are involved in recessive cone dystrophy (Ba-Abbad et al., 2016; Wycisk et al., 2006a). Mice with a premature stop mutation in *Cacna2d4* show abnormal morphology of ribbon synapses in the rods and cones (Wycisk et al., 2006b). In mice loss of *Cacna2d4* abolished rod synaptogenesis and synaptic transmission and disrupted mGluR6 clustering (Y. Wang et al., 2017). This suggests that CACN2D4 is involved in visual perception by the eye, but it does not provide an argument for its involvement in higher order cortical specialisation and visual perception. There was no indication that the family members with developmental colour agnosia suffered from an ophthalmic problem. This may also suggest that the splice-site mutation that we identified is not deleterious. Furthermore, *CACNA2D4* has been implicated in autism spectrum disorders (X. Liao, Liao, & Li, 2020), and a deletion in the *CACNA2D4* gene was found in a patient with bipolar disorder, but it is unknown if this patient suffers with vision problems (Van Den Bossche et al., 2012). Our attempts to trace this patient for developmental colour agnosia testing failed. Studies of other CACNA subtypes showed that thermal pain and tactile stimulation in the *Cacna2d*3 mutant mice triggered strong crossactivation of brain regions involved in vision, olfaction, and hearing. This study suggested that this knockout model could be a model for synaesthesia (Neely et al., 2010), which is a disorder related to developmental colour agnosia (Rich & Mattingley, 2002).

*MYO15A* mutations are associated with deafness due to its critical role for the formation of stereocilia in cochlear hair cells (Probst et al., 1998). Recessive mutations lead to autosomal recessive nonsyndromic hearing loss (A. Wang et al., 1998). *MYO15A* mutations are associated with the Usher syndrome having affected hearing (Baux et al., 2017). Visual perception is also affected In Usher syndrome from the age of 10 years onwards. Vision loss is caused by retinopathia which is due to degeneration of retina cells, usually the rods cells.

Since no direct link to visual perception was found for the other genes, their possible function in the (developing) brain was considered for these genes. It has been indicated that glutamate receptor, ionotropic, N-methyl D-aspartate-associated protein 1 (glutamate binding) (*GRINA*) is involved in nervous system development and disease mainly through signalling mechanisms of neuronal survival (Chen, Yang, Lai, Liu, & Zhu, 2020). Upregulation of *GRINA* transcripts was observed in post-mortem brains of patients with major depressive disorder, supporting a role in brain functions (Goswami et al., 2013). *Grina* knock-out (KO) mice do not display an obvious phenotype (Nielsen et al., 2011). This KO study showed that the effects of the *GRINA* gene are probably subtle. Since it is likely that effects on higher order visual perception will not be evident in mice, *GRINA* remains a potentially interesting candidate gene for consideration of a role in developmental colour agnosia. The effect of the mutation is probably damaging or deleterious (SIFT) and the site of mutation is highly conserved for nucleotide sequence and amino acid motif.

*DDX25 (DEAD (Asp-Glu-Ala-Asp) box polypeptide 25)* encodes a member of the large DEAD-box RNA helicases. Members of this protein family share high structural conservation and are generally involved in developmental processes such as gametogenesis, germ cell specification, and stem cell biology. Data on roles of DDX proteins in the brain are scarce. However, recently two members have been implicated in brain processes. Firstly, a homozygous frameshift deletion in *DDX59* (c.185del: p.Phe62fs*13) has been reported in a family presenting with orofaciodigital syndrome phenotype associated with a broad neurological involvement characterized by microcephaly, intellectual disability, epilepsy, and white matter signal abnormalities associated with cortical and subcortical ischemic events (Salpietro et al., 2018). Further research in *Drosphila* showed impaired development of peripheral and central nervous system upon loss-of-function of *DDX59*, supporting a conserved role of this DEAD-box RNA helicase in neurological function (Salpietro et al., 2018). Secondly, mutations in *DDX3X* cause neurodevelopmental abnormalities in females, including intellectual disability, cortical malformations, autism and epilepsy (Lennox et al., 2020). In this study it was shown that in vivo depletion of Ddx3x in mice reduced neuronal differentiation and impaired corticogenesis (Lennox et al., 2020).

A role of DDX25 in the brain has not been studied. Sofar *DDX25* has been implicated in spermatogenesis and primordial germ cell development (Kavarthapu et al., 2019; Tsai-Morris, Sheng, Lee, Lei, & Dufau, 2004). Polymorphisms in the *DDX25* gene have been found that alter protein function (Tsai-Morris et al., 2007). During our expression analyses of DDX25 we found significant expression in the brain. The Allen Brain Atlas Data Portal (http://casestudies.brain-map.org/celltax#section_explorea) showed expression of *Ddx25* in mouse cerebral cortex, nucleus accumbens, several thalamic and hypothalamic nuclei, locus coeruleus, raphe nucleus, inferior olivary complex, and absence in globus pallidus, and most of the midbrain and brain stem. Further exploration of mouse cortical gene expression of the data of (Tasic, Menon, Nguyen, Kim, Jarsky, Yao, Levi, Gray, Sorensen, Dolbeare, Bertagnolli, Goldy, Shapovalova, Parry, Lee, Smith, Bernard, Madisen, Sunkin, Hawrylycz, Koch, & Zeng, 2016) through the Single Cell Portal (https://portals.broadinstitute.org/single_cell) showed expression of *Ddx25* in all subtypes of interneurons and pyramidal cells. Transcripts were absent in glial cells (not shown). Mining of more recent data from single-cell transcriptomics (Hodge et al., 2019) showed that expression of *DDX25* in human and mouse cortex is relatively low and is predominating in somatostatin- and parvalbumin-expressing interneurons, and in several types of pyramidal neurons, mostly located in layers 5-6 (dataset human M1; Fig 4A). In the dataset ‘Multiple cortical areas’ no evident expression was detected. There was no clear age-related change in human brain samples (Fig 3). In mouse single-cell data sets there was very low expression in parvalbumin-expressing interneurons and layer 5 pyramidal neurons (REFs; Fig 4B). Overall, *DDX25* appears a gene with low and restricted expression in the adult cortex. Since it is part of a little-studied family causing severe neurological problems, as in the case of *DDX3A* and *DDX59* mutations, *DDX25* should still be considered a potential candidate gene for involvement in developmental colour agnosia.

*Staufen double-stranded RNA binding protein 1* gene *STAU1* carried a possibly damaging (Polyphen) and tolerated (SIFT) mutation in a highly conserved amino acid motif. Staufen1 is a key protein involved in the local translation of mRNA in the dendrite and is involved in protein synaptic transmisson. Cultured neurons from *Stau1* knockout mice showed reduced dendritic trees and developed fewer synapses, but the mice did not show any obvious defects in brain development (Vessey et al., 2008). We note that recently a molecular and functional relationship between Stau1 and DEAD-box RNA helicases (Ddx5/17) was established in a complex with the Kruppel-like factor Klf4 (Moon et al., 2018). It was shown that this Klf4/Stau1/Ddx5/7 protein complex regulates neurogenesis-associated mRNAs and plays a role in mammalian corticogenesis. Furthermore, local regulatory networks containing *STAU1* have been identified in cognitive decline in older adults (Tasaki et al., 2018). In accord with a neuronal role, *STAU1* appeared expressed in virtually all types of cortical neurons in mouse and man, but not in non-neuronal cells of the cerebral cortex.

The *Mastermind-like 2* (*MAML2*) gene, encoding a transcriptional activator of the Notch signalling pathway (Ribeiro & Wallberg, 2009), has been implicated in cancers, but has no known function in the nervous system. The effect of the found mutation is probably damaging in Polyphen and tolerated in SIFT. The nucleotide and amino acid sequence is moderate and highly conserved, respectively. In this study *MAML2* is notable since its brain expression is increased during development (age 0.1-0.5 years) in comparison to older ages (Fig 3). Interestingly, it displayed highest expression by several non-neuronal cell types. In the human cortex these were: oligodendrocyte progenitor cell L1-6 PDGFRA COL20A1 (trimmed mean expression level 10.2), several types of astrocytes (level 9.8-10.3) and microglia (level 10.9) (Fig 4). Neuronal expression of MAML2 at appreciable level occurred in three subtype of inhibitory neurons: inhibitory neuron L1 Pax6 Mir 101-1 (level: 8.2), inhibitory neuron L1-2 VIP WNT4 (level: 8.54), inhibitory neuron L1-6 SST NPY (level 8.67). Two excitatory cell types expressed MAML2: L3 LINC00507 and L3 FEZF2 (respectively at level 8.58 and 7.08). In a single-dataset of the mouse cortex and hippocampus, *Maml2* gene was highest expressed in cells of the dentate gyrus and CA region (Fig 4). The Allen Mouse Brain Atlas confirmed the selective high expression of *Maml2* in the dentate gyrus.

The expression of *transmembrane emp24 protein transport domain containing 3* (*TMED3*) is noticeable in its increased between the age of 0.3-8 years in the human prefrontal cortex (fig 3). *TMED3* expression is assigned to all interneurons and pyramidal cells in the mouse cortex (Single Cell Portal;(Tasic, Menon, Nguyen, Kim, Jarsky, Yao, Levi, Gray, Sorensen, Dolbeare, Bertagnolli, Goldy, Shapovalova, Parry, Lee, Smith, Bernard, Madisen, Sunkin, Hawrylycz, Koch, & zheng, 2016), but displays very low to no expression in more recent human and mouse datasets. TMED3 participates in tumor progression, but functions of TMED3 in the brain have not been described. Interestingly, it has been proposed that in tumors TMED3 promotes endogenous Wnt-Tcf activity, which is an important pathway in brain development, and Il11/Stat3 signalling (Duquet et al., 2014; Zheng et al., 2016).development. The effect of the mutation is possibly damaging in Polyphen and tolerated in SIFT and conservation is moderate for nucleotide and highly conserved for amino acid sequence (Table 3).

The effect of the mutation found in the *DEPD*C *(DEP domain-containing 7)* gene is a possibly damaging one in Polyphen and tolerated in SIFT (Table 3). Brain expression of *DEPDC7* was not detectable. However, it has been suggested that *DEPDC7* DNA hypomethylation may be associated with depression (Córdova-Palomera et al., 2015). The biological function of the protein encoded by the *DEPDC7* (DEP domain-containing 7) gene is poorly understood. The DEPDC7 protein participates in gene regulation by NF-κB (D’Andrea et al., 2014). It has been shown that it inhibits tumor growth (Liao, Wang, Wang, Li, & Lin, 2017). The gene has been detected amongst nine genes in a microdeletion in a patient with cryptorchidism and azoospermia (Seabra et al., 2014).

No data are available on the function of olfactory receptors *OR1J1 (olfactory receptor, family 1, subfamily J, member 1)* and *OR8D4*. Expression of ORs has been observed in other tissues than olfactory neurons. *OR8D4* has been encountered as a risk gene of systemic lupus erythematosus. The mutation for *OR1J1* is benign for Polyphen and tolerated for SIFT. The mutation in *OR8D4* is benign in Polyphen and deleterious in SIFT.

*RABEPK (Rab9 effector protein with Kelch motifs)* has not been studied to any extent. It is thought to participate in protein transport in the trans-Golgi network. The effect of the mutations is a probably damaging in Polyphen and deleterious in SIFT and the conservation is high for both sequences. Expression of the RABEPK gene is not detectable in single-cell data sets of the human cortex and in very low level in several types of interneurons and pyramidal cells in the mouse cortex and hippocampus (Fig 4).

Taken together, this study is the first attempt to pinpoint genomic regions and genetic variants in developmental colour agnosia, a cognitive trait affecting the understanding of colours. This study was made possible by the finding and characterization of an unique family carrying this trait (Nijboer, van Zandvoort, et al., 2007b; van Zandvoort et al., 2007). We assumed that developmental colour agnosia is an extremely rare trait. Additional subjects were not found since initiation of this study in 2012. Developmental colour agnosia is not a disorder that requires medical or psychological attention, and will hence stay unnoticed, which hampers the estimation of prevalence.

In addition to number of subjects, this study is also limited by the assumption that developmental colour agnosia is inherited in an autosomal dominant manner with complete penetrance, based on the segregation pattern in the family. Furthermore, the exomes were analysed in 2012 and since then the quality of capture and sequencing have improved. In addition, we cannot exclude the involvement of genomic rearrangement or mutations in noncoding parts of the genome which we would not have captured using exome sequencing. Within these limitations this study indicates 11 genes that link to the trait. Amongst these there are candidate that have been implicated in neural processes and functions (*CACNA2D4, MYO15A, DDX25, GRINA*), that have an indirect link to brain function or development (*STAU1, MAML2, TMED3*), while others remain unlikely due to lack of brain expression, involvement of nonneural traits (*OR1J1, OR8D4, DEPDC7, RABEPK*)

In summary, we identified 19 genomic regions and 11 coding variants in hereditary colour agnosia by exome sequencing. Since replication is not possible yet due to the uniqueness of this identified family we cannot pinpoint the causal variant at this stage. Further research is required to identify the genetic origins of developmental colour agnosia in this family.

## Data Availability

Data can be obtained from the first authors upon request

## Acknowledgements

This project was supported financially by the Neuroscience Center Utrecht as part of the Focus & Massa programme of the Utrecht University.

## Author contributions

Conceptualization: MJvZ, TCN, and JPHB; Cohort assembly: TCN, MJvZ, EHB; Experiments: EVSH, GvH, Computational analyses: EVSH, GvH, CdK, BPCK, PvdS, CT, BvdZ; Data interpretation: EVSH, GvH, JPHB; Writing original draft, TCN, EVSH, GvH.; Writing – review & editing: TCH, JPHB, GvH.

